# A functional *TGFB1* polymorphism in the donor associates with long-term graft survival after kidney transplantation

**DOI:** 10.1101/2021.07.05.21260045

**Authors:** Felix Poppelaars, Mariana Gaya da Costa, Bernardo Faria, Siawosh K. Eskandari, Jeffrey Damman, Marc A. Seelen

## Abstract

**Introduction:** Improvement of long-term outcomes in kidney transplantation remains one of the most pressing challenges, yet drug development is stagnating. Human genetics offers an opportunity for much-needed target validation in transplantation. Conflicting data exist about the effect of transforming growth factor-beta 1 (TGF-β1) on kidney transplant survival since TGF-β1 has pro-fibrotic and protective effects. We investigated the impact of a recently discovered functional *TGBF1* polymorphism on kidney graft survival.

**Methods:** We performed an observational cohort study analyzing recipient and donor DNA in 1,271 kidney transplant-pairs from the University Medical Center Groningen in The Netherlands and associated a low-producing *TGBF1* polymorphism (rs1800472-C>T) with 5, 10, and 15-year death-censored kidney graft survival.

**Results:** Donor genotype frequencies of rs1800472 in *TGBF1* differed significantly between patients with and without graft loss (*P*=0.014). Additionally, the low-producing *TGBF1* polymorphism in the donor was associated with an increased risk of graft loss following kidney transplantation (HR 2.12 for the T-allele; 95%-CI 1.18–3.79; *P*=0.012). The incidence of graft loss within 15 years of follow-up was 16.4% in the CC-genotype group and 31.6% in the CT-genotype group. After adjustment for transplant-related covariates, the association between the *TGBF1* polymorphism in the donor and graft loss remained significant. In contrast, there was no association between the *TGBF1* polymorphism in the recipient and graft loss.

**Conclusion:** Kidney allografts possessing a low-producing *TGBF1* polymorphism have a higher risk of late graft loss. Our study adds to a growing body of evidence that TGF-β1 is beneficial, rather than harmful, for kidney transplant survival.

## Introduction

Short-term outcomes following kidney transplantation have dramatically improved in the past 25 years, adding over one million life-years to patients in the United States alone. Nonetheless, improving the long-term transplant outcomes remains a crucial challenge.(1) The alloimmune response is recognized as a major contributor to late kidney transplant failure.(2) Furthermore, cytokines play a pivotal role in orchestrating the immune response.(3) Understanding the contribution of cytokines to donor-recipient incompatibility in kidney transplantation, therefore, is crucial as it can lead to the development of novel treatment strategies. Transplantation is a unique situation from a genetic and an immunological perspective, as two genomes are brought together. Genetic differences between the donor and recipient subsequently lead to immunological injury.(4) Although the recipient primarily drives the alloimmune response, the release of inflammatory triggers by the donor kidney is gaining traction as an essential additional mechanism.(5) Specifically, recent studies indicate that the local inflammatory response by the donor kidney significantly impacts transplant outcome.(6)

Among all cytokines, transforming growth factor-beta (TGF-β) is a multifaceted and -functional cytokine that is synthesized by nearly every cell type.(7) To date, three main isoforms of TGF-β have been identified in humans, (that is TGF-β1, TGF-β2, and TGF-β3), which are encoded by distinct genes (*TGFB1, TGFB2*, and *TGFB3* respectively). Between these, TGF-β1 is the most common and best-characterized isoform. The functions of TGF-β1 range from regulating cellular processes (such as differentiation, migration, and apoptosis) to initiating the production of extracellular matrix proteins.(8) Despite this complexity, the TGF-β signaling pathway relies on a simple ligand-activated receptor complex. More specifically, signaling is initiated when dimerized TGF-β1 binds surface-tethered TGF-β receptors, namely TGF-βR1 and TGF-βR2 (Fig. 1A).(9) This binding activates TGF-βR2, allowing it to phosphorylate TGF-βR1, which then propagates the signal intracytoplasmically by phosphorylating transcription factors of the small mothers against decapentaplegic homolog (SMAD) family, SMAD2 and SMAD3 (Fig. 1B).(9) Upon phosphorylation, SMAD2 and -3 trimerize with an obligate partner, SMAD4, permitting the nuclear translocation of the complex and, with the help of nuclear cofactors, transcription of TGF-β target genes (Fig. 1B).(9)

**Figure 1.**
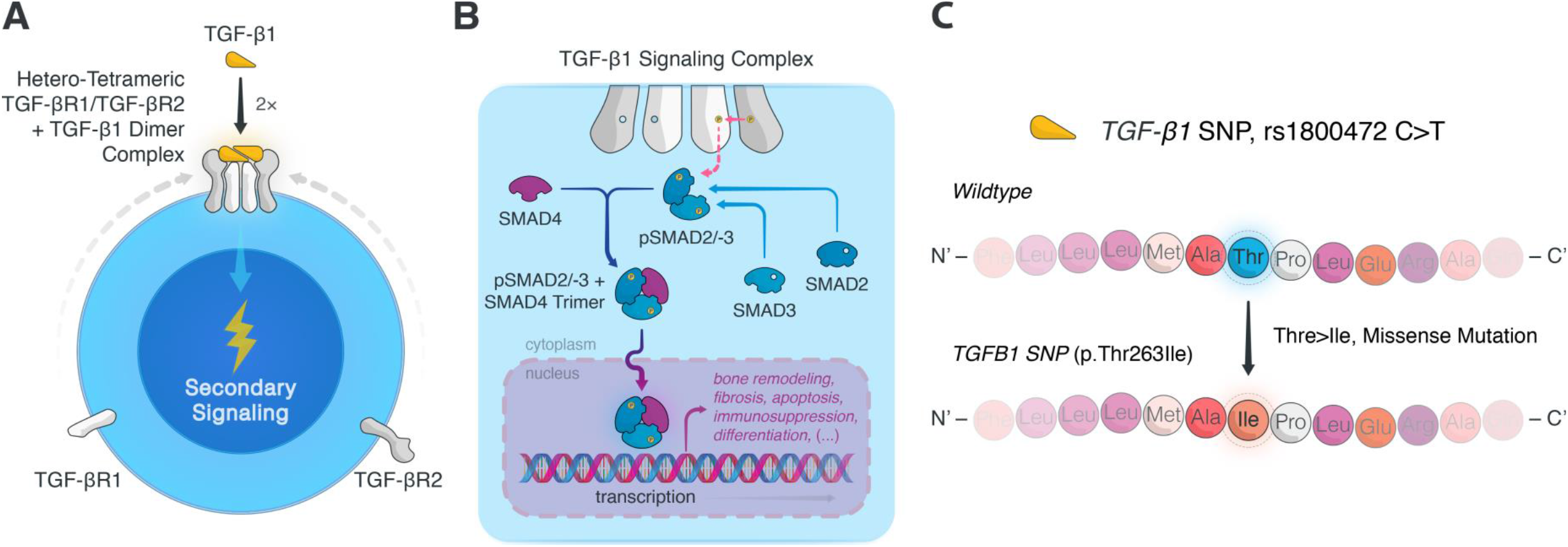
TGF-β1 signaling pathway and examined TGFB1 Thr263Ile gene variant. (**A**) TGF-β1 signaling occurs when TGF-β1 forms a complex with surface-bound TGF-β receptors 1 and 2 (TGF-βR1 and TGF-βR2 respectively). Specifically, two heterodimers of TGF-βR1/TGF-βR2 coalesce in the presence of dimeric TGF-β1, resulting in hetero-tetrameric complex. (**B**) The proximity of the intracytoplasmic tails of TGF-βR2 initiates the sequential phosphorylation of TGF-βR1 and the SMAD signal transducers, SMAD2 and -3. (Of note, although it is uncommon, following TGF-βR1 phosphorylation non-SMAD-mediated signaling can also occur.) Once SMAD2/-3 has formed a dimeric unit, it can bind SMAD4, leading to nuclear translocation of the trimeric pSMAD2/-3 and SMAD4 complex. Inside the nucleus, the trimeric complex elicits transcription of TGF-β target genes, resulting in a myriad of cellular responses from bone remodeling to fibrosis, to apoptosis and immunosuppression. (**C**) To appreciate the potential role of *TGFB1*-related SNPs in kidney transplant recipients and donor transplant kidneys, we assessed the association between transplant survival outcomes and the *TGFB1* SNP rs1800472C>T, which causes a missense mutation (p.Thr263Ile) in TGF-β1. Ile, isoleucine; SMAD, Small mothers against decapentaplegic homologue; TGF-β, transforming growth factor beta; TGF-βR, transforming growth factor beta receptor; Thr, threonine.

Among its many biological roles, TGF-β1 is predominantly known for being a critical driver of fibrosis in various diseases and conditions.(10) As a result, modulation of TGF-β1 activation and signaling is currently pursued as a therapeutic strategy to halt cancer progression as well as to prevent fibrosis after surgery and in chronic diseases.(11) In recent years, however, the protective functions of TGF-β1 have attracted much attention and are now deemed equally important. Evidence from animal models and *in vitro* experiments demonstrate that the protective effects of TGF-β1 range from inhibiting inflammation to inducing autophagy.(7) Accordingly, the role of TGF-β1 in disease is context-depending and may be protective or harmful.

In kidney transplantation, TGF-β1 has been a topic of interest for many years and TGF-β1 has been suggested to impact allograft survival in different ways.(12) Initially, multiple reports showed upregulation of TGF-β1 expression and signaling in kidney transplants during rejection.(13,14) Separately, plasma levels of TGF-β1 were shown to be a potential biomarker of progressive chronic kidney disease in certain populations.(15) Animal studies then demonstrated that TGF-β1 overexpression in the kidney induced interstitial proliferation, tubular autophagy, and fibrosis.(16) In contrast, genetic deficiency of TGF-β1 in mice leads to multiorgan inflammation (including that of the kidney).(17) In the context of these findings, Du and colleagues found that TGF-β1 plasma levels were positively associated with long-term graft survival in kidney transplant recipients.(18) Finally, local TGF-β1 expression in the kidney allograft during rejection has been associated with a favorable outcome.(19,20) In these studies, however, it often remains unclear whether the association found with TGF-β1 is a cause or consequence of the pathology. Genetic studies have therefore used single nucleotide polymorphisms (SNPs) in the Transforming Growth Factor Beta 1 gene (*TGFB1*) to dissect the impact of TGF-β1 signaling on kidney transplant outcome. However, these studies have primarily been retrospective and underpowered, and, thus, they are often inconclusive in their analyses.

To elucidate the current conflicting data, we investigated the impact of a polymorphism in *TGFB1* on long-term outcomes in kidney transplantation patients as a model for target validation (Fig. 1C). We specifically chose to study the *TGFB1* Thr263Ile variant (rs1800472 C>T) since it was identified as a major genetic driver of plasma TGF-β1 levels in a recent study by Höglund and colleagues using whole-genome sequencing data. In their study, the minor allele (=T-allele) of this polymorphism was shown to be significantly associated with lower plasma levels of TGF-β1.(21) Furthermore, the overall heritability of the differences observed in the plasma TGF-β1 concentration was 22.9%, while the heritability conditioned of this variant alone was 16.3%. We evaluated in the present study the association between this recently discovered low-producing *TGFB1* polymorphism and long-term kidney graft survival. Additionally, our secondary endpoints were delayed graft function (DGF) and biopsy-proven acute rejection (BPAR).

## Materials and Methods

### Patient selection and study end-point

The primary endpoint in this study was death-censored graft survival, defined as the need for dialysis or re-transplantation. Secondary endpoints were delayed graft function (DGF, defined by the United Network for Organ Sharing as “*the need for at least one dialysis treatment in the first week after kidney transplantation*”)(22) and biopsy-proven acute rejection (BPAR, according to the Banff 2007 classification). We enrolled patients who underwent single kidney transplantation at the University Medical Center Groningen in the Netherlands between 1993 and 2008. From the 1430 kidney transplantations, 1271 recipient and donor pairs were included in the cohort as previously described.(23,24) Subjects were excluded due to technical complications during surgery, lack of DNA, re-transplantation, or loss of follow-up. This study was performed in accordance with the declaration of Helsinki and all patients provided written informed consent. The medical ethics committee of the University Medical Center Groningen approved the study under file n° METc 2014/077.

### DNA extraction and *TGFB1* genotyping

Peripheral blood mononuclear cells were isolated from blood or splenocytes collected from the recipients and donors. DNA was extracted with a commercial kit as per the manufacturer’s instructions and stored at -80°C. Genotyping of the SNPs was determined via the Illumina VeraCode GoldenGate Assay kit (Illumina, San Diego, CA, USA), according to the manufacturer’s instructions. Genotype clustering and calling were performed using BeadStudio Software (Illumina). The overall genotype success rate was 99.5% and six samples with a high missing call rate were excluded from subsequent analyses.

### Statistical analysis

Statistical analyses were performed using SPSS software version 25 (SPSS Inc, Chicago, IL, USA). Data are displayed as mean ± standard deviation for parametric variables, median [IQR] for non-parametric variables, and nominal data as the total number of patients and the percentage [n (%)]. Differences between groups were examined with the Student’s t-test for normally distributed variables or the Mann-Whitney-U test for the not-normally distributed variables, and χ2 test for categorical variables, respectively. Log-rank tests were performed between different genotypes to assess the difference in the incidence of graft loss. Univariable analysis was performed to determine the association of genetic, donor, recipient, and transplant characteristics with graft survival. The factors identified in these analyses were thereafter tested in a multivariable Cox regression. Additionally, multivariable Cox regression with a stepwise forward selection was performed. Tests were two-tailed and regarded as statistically significant when *P*<0.05.

## Results

### Patient characteristics and long-term graft survival

Baseline demographics and clinical characteristics of the 1,271 kidney transplant donor-recipient pairs are shown in Table 1. The mean follow-up after transplantation was 6.16 years ± 4.21 with a maximum follow-up period of 15 years. During follow-up, 215 grafts (16.9%) were lost, and the causes of graft failure included rejection (n = 126, including acute rejection, chronic antibody-mediated rejection and transplant glomerulopathy), vascular causes (n = 12), recurrence of primary disease (n = 16), surgical complications (n = 33), other causes (n = 16), or unknown (n = 12). The following characteristics were significantly associated with graft loss in univariate analysis; donor age, donor blood type (ABO *vs* others), donor type (living *vs* cadaveric), recipient age, recipient blood type (ABO *vs* others), use of cyclosporin, use of corticosteroids, cold ischemia time, warm ischemia time, and DGF.

**Table 1:**
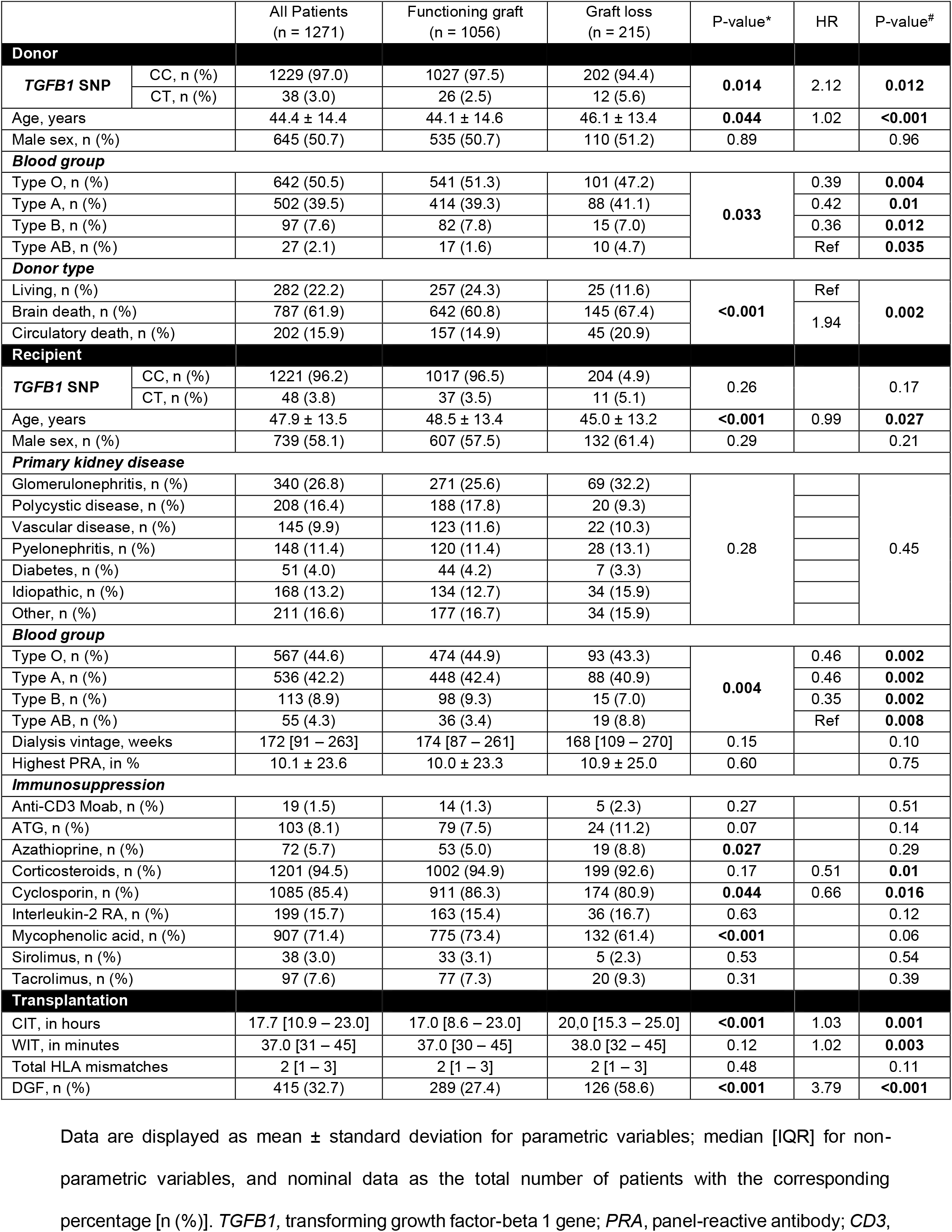
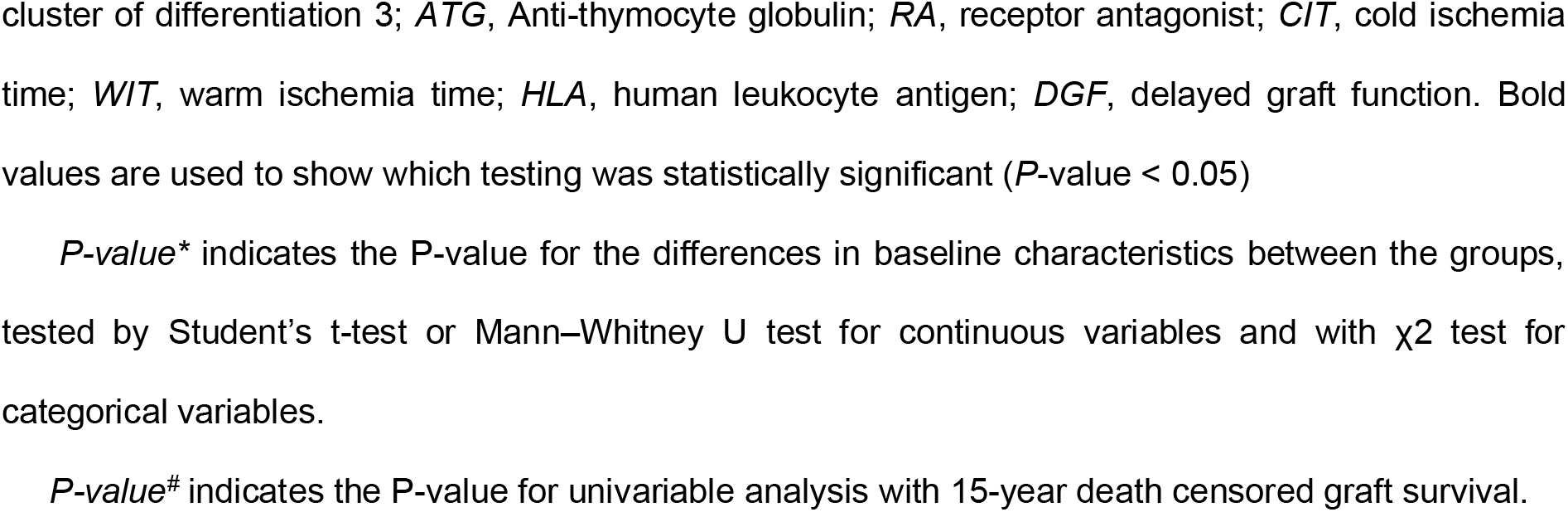
Baseline characteristics of the donors and recipients.

|  |  | All Patients<br>(n = 1271) | Functioning graft<br>(n = 1056) | Graft loss<br>(n = 215) | P-value* | HR | P-value# |
| --- | --- | --- | --- | --- | --- | --- | --- |
| Donor |  |  |  |  |  |  |  |
| TGFB1 SNP | CC, n (%) | 1229 (97.0) | 1027 (97.5) | 202 (94.4) | 0.014 | 2.12 | 0.012 |
|  | CT, n (%) | 38 (3.0) | 26 (2.5) | 12 (5.6) |  |  |  |
| Age, years |  | 44.4 ± 14.4 | 44.1 ± 14.6 | 46.1 ± 13.4 | 0.044 | 1.02 | <0.001 |
| Male sex, n (%) |  | 645 (50.7) | 535 (50.7) | 110 (51.2) | 0.89 |  | 0.96 |
| Blood group |  |  |  |  |  |  |  |
| Type O, n (%) |  | 642 (50.5) | 541 (51.3) | 101 (47.2) | 0.033 | 0.39 | 0.004 |
| Type A, n (%) |  | 502 (39.5) | 414 (39.3) | 88 (41.1) |  | 0.42 | 0.01 |
| Type B, n (%) |  | 97 (7.6) | 82 (7.8) | 15 (7.0) |  | 0.36 | 0.012 |
| Type AB, n (%) |  | 27 (2.1) | 17 (1.6) | 10 (4.7) |  | Ref | 0.035 |
| Donor type |  |  |  |  |  |  |  |
| Living, n (%) |  | 282 (22.2) | 257 (24.3) | 25 (11.6) | <0.001 | Ref | 0.002 |
| Brain death, n (%) |  | 787 (61.9) | 642 (60.8) | 145 (67.4) |  | 1.94 |  |
| Circulatory death, n (%) |  | 202 (15.9) | 157 (14.9) | 45 (20.9) |  |  |  |
| Recipient |  |  |  |  |  |  |  |
| TGFB1 SNP | CC, n (%) | 1221 (96.2) | 1017 (96.5) | 204 (4.9) | 0.26 |  | 0.17 |
|  | CT, n (%) | 48 (3.8) | 37 (3.5) | 11 (5.1) |  |  |  |
| Age, years |  | 47.9 ± 13.5 | 48.5 ± 13.4 | 45.0 ± 13.2 | <0.001 | 0.99 | 0.027 |
| Male sex, n (%) |  | 739 (58.1) | 607 (57.5) | 132 (61.4) | 0.29 |  | 0.21 |
| Primary kidney disease |  |  |  |  |  |  |  |
| Glomerulonephritis, n (%) |  | 340 (26.8) | 271 (25.6) | 69 (32.2) | 0.28 |  | 0.45 |
| Polycystic disease, n (%) |  | 208 (16.4) | 188 (17.8) | 20 (9.3) |  |  |  |
| Vascular disease, n (%) |  | 145 (9.9) | 123 (11.6) | 22 (10.3) |  |  |  |
| Pyelonephritis, n (%) |  | 148 (11.4) | 120 (11.4) | 28 (13.1) |  |  |  |
| Diabetes, n (%) |  | 51 (4.0) | 44 (4.2) | 7 (3.3) |  |  |  |
| Idiopathic, n (%) |  | 168 (13.2) | 134 (12.7) | 34 (15.9) |  |  |  |
| Other, n (%) |  | 211 (16.6) | 177 (16.7) | 34 (15.9) |  |  |  |
| Blood group |  |  |  |  |  |  |  |
| Type O, n (%) |  | 567 (44.6) | 474 (44.9) | 93 (43.3) | 0.004 | 0.46 | 0.002 |
| Type A, n (%) |  | 536 (42.2) | 448 (42.4) | 88 (40.9) |  | 0.46 | 0.002 |
| Type B, n (%) |  | 113 (8.9) | 98 (9.3) | 15 (7.0) |  | 0.35 | 0.002 |
| Type AB, n (%) |  | 55 (4.3) | 36 (3.4) | 19 (8.8) |  | Ref | 0.008 |
| Dialysis vintage, weeks |  | 172 [91 – 263] | 174 [87 – 261] | 168 [109 – 270] | 0.15 |  | 0.10 |
| Highest PRA, in % |  | 10.1 ± 23.6 | 10.0 ± 23.3 | 10.9 ± 25.0 | 0.60 |  | 0.75 |
| Immunosuppression |  |  |  |  |  |  |  |
| Anti-CD3 Moab, n (%) |  | 19 (1.5) | 14 (1.3) | 5 (2.3) | 0.27 |  | 0.51 |
| ATG, n (%) |  | 103 (8.1) | 79 (7.5) | 24 (11.2) | 0.07 |  | 0.14 |
| Azathioprine, n (%) |  | 72 (5.7) | 53 (5.0) | 19 (8.8) | 0.027 |  | 0.29 |
| Corticosteroids, n (%) |  | 1201 (94.5) | 1002 (94.9) | 199 (92.6) | 0.17 | 0.51 | 0.01 |
| Cyclosporin, n (%) |  | 1085 (85.4) | 911 (86.3) | 174 (80.9) | 0.044 | 0.66 | 0.016 |
| Interleukin-2 RA, n (%) |  | 199 (15.7) | 163 (15.4) | 36 (16.7) | 0.63 |  | 0.12 |
| Mycophenolic acid, n (%) |  | 907 (71.4) | 775 (73.4) | 132 (61.4) | <0.001 |  | 0.06 |
| Sirolimus, n (%) |  | 38 (3.0) | 33 (3.1) | 5 (2.3) | 0.53 |  | 0.54 |
| Tacrolimus, n (%) |  | 97 (7.6) | 77 (7.3) | 20 (9.3) | 0.31 |  | 0.39 |
| Transplantation |  |  |  |  |  |  |  |
| CIT, in hours |  | 17.7 [10.9 – 23.0] | 17.0 [8.6 – 23.0] | 20.0 [15.3 – 25.0] | <0.001 | 1.03 | 0.001 |
| WIT, in minutes |  | 37.0 [31 – 45] | 37.0 [30 – 45] | 38.0 [32 – 45] | 0.12 | 1.02 | 0.003 |
| Total HLA mismatches |  | 2 [1 – 3] | 2 [1 – 3] | 2 [1 – 3] | 0.48 |  | 0.11 |
| DGF, n (%) |  | 415 (32.7) | 289 (27.4) | 126 (58.6) | <0.001 | 3.79 | <0.001 |

3 Data are displayed as mean ± standard deviation for parametric variables; median [IQR] for non-
4 parametric variables, and nominal data as the total number of patients with the corresponding
5 percentage [n (%)]. *TGFB1*, transforming growth factor-beta 1 gene; *PRA*, panel-reactive antibody; *CD3*,
cluster of differentiation 3; *ATG*, Anti-thymocyte globulin; *RA*, receptor antagonist; *CIT*, cold ischemia time; *WIT*, warm ischemia time; *HLA*, human leukocyte antigen; *DGF*, delayed graft function. Bold values are used to show which testing was statistically significant ( $P$ -value < 0.05)
*P-value*\* indicates the P-value for the differences in baseline characteristics between the groups, tested by Student's t-test or Mann–Whitney U test for continuous variables and with $\chi^2$ test for categorical variables.
*P-value*<sup>#</sup> indicates the P-value for univariable analysis with 15-year death censored graft survival.

### Distribution of the *TGFB1* genetic variant

The observed genotypic frequencies of the Thr263Ile *TGFB1* variant (rs1800472 C>T) did not significantly differ between recipients (n = 1269; CC, 96.2%; CT, 3.8%; TT, 0%) and donors (n = 1267; CC, 97.0%; CT, 3.0%; TT, 0%) (*P* = 0.55). No homozygosity was observed for this *TGFB1* polymorphism, but the distribution of the SNP was in Hardy−Weinberg equilibrium. The genotypic frequencies of the *TGFB1* polymorphism in donors and recipients were significantly higher than those reported by the 1000 genomes project (*P* = 0.013), but not compared to their European cohort (*P* = 0.60).(25) The proportion of grafts with DGF significantly differed based on recipient *TGFB1* genotype (45.8% in CT vs. 32.2% in CC, *P* = 0.048), but not for donor *TGFB1* genotype (36.8% in CT vs. 32.5% in CC, *P* = 0.58) (Supplementary Data). In logistic regression, recipients carrying the T-allele of the *TGFB1* variant showed a trend towards a higher risk of DGF (OR = 1.78 compared to C-allele; 95%-CI: 1.00 – 3.19; *P* = 0.051). There was no difference in the overall BPAR frequency between the *TGFB1* genotypes in the donor (33.9% in CT vs. 34.2% in CC, *P* = 0.97) or the recipient (25.0% in CT vs. 34.4% in CC, *P* = 0.18) (Supplementary Data). By contrast, the distribution of the *TGFB1* polymorphism in the donor, but not the recipient, differed significantly between patients with and without graft loss after complete follow-up (Table 1, *P* = 0.014). More specifically, the T-allele of the *TGFB1* SNP was more prevalent in kidney grafts that were lost during the follow-up period. These data suggest that TGF-β1 expression by the donor kidney might impact long-term graft survival in kidney transplantation.

### Long-term kidney graft survival according to the *TGFB1* genotypes

Kaplan–Meier survival analysis showed that the *TGFB1* SNP in the donor was associated with an increased risk for graft loss during follow-up (Figure 2). The *TGFB1* variant in the donor was significantly associated with 10-, and 15-year death-censored kidney graft survival in Kaplan–Meier survival analyses (Fig. 2B – C), but not with 5-year graft survival (Fig. 2A). After complete follow-up, the incidence of graft loss was 16.4% in the reference CC-genotype group and 31.6% in the CT-genotype group, respectively. The *TGFB1* variant in the recipient was not associated with death-censored kidney graft survival in Kaplan–Meier survival analyses (Fig. 2D – F). Subgroup analysis for recipient sex and donor type did not change these results (Supplementary Data).

**Figure 2.**
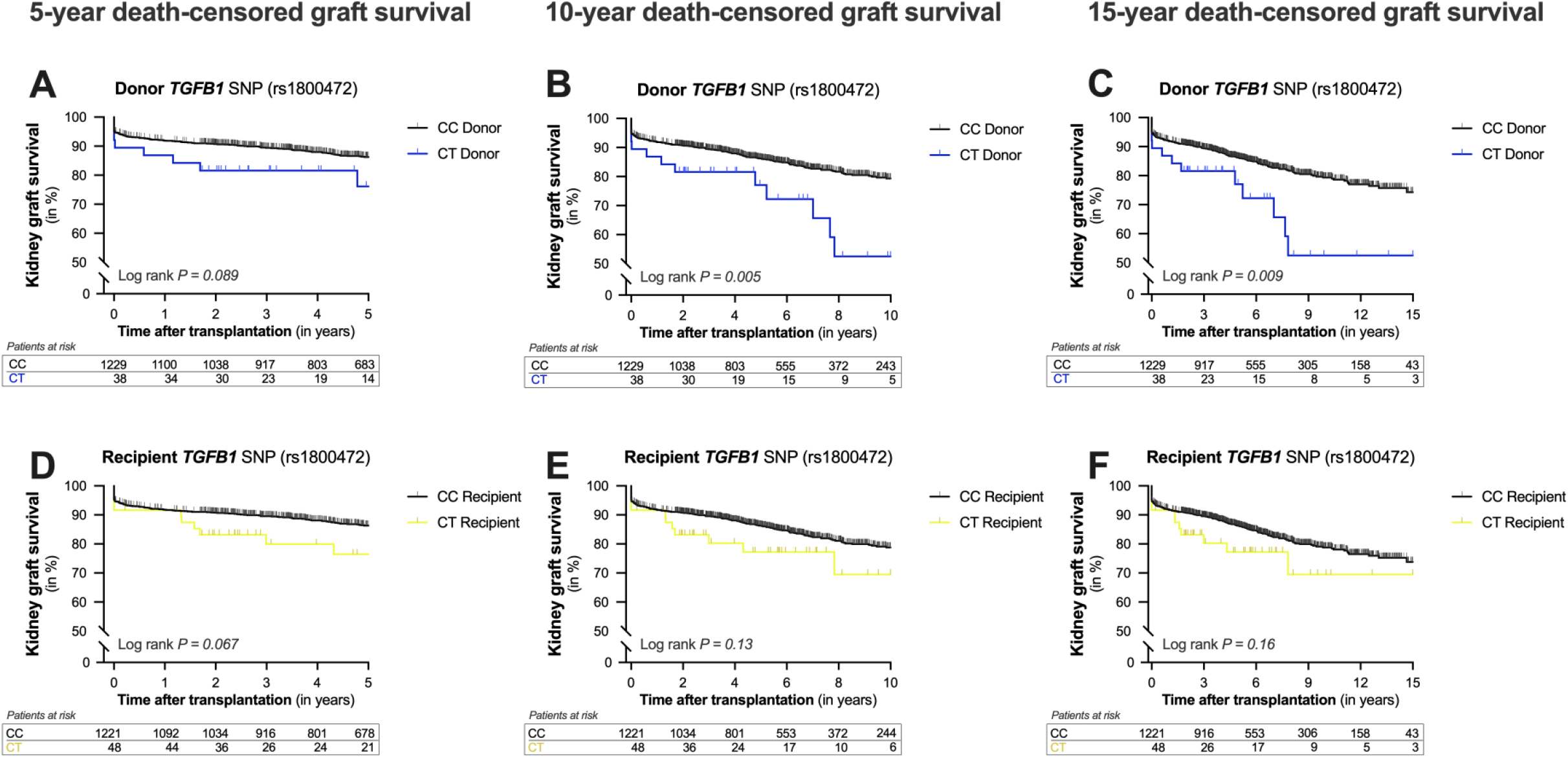
Kaplan-Meier curves for 5, 10, 15-year death-censored graft survival after kidney transplantation according to the presence of the *TGFB1* variant in the donor and recipient. Cumulative 5- (**A, D**), 10- (**B, E**), and 15-year (**C, F)** death-censored kidney graft survival according to the presence of the Thr263Ile variant in the transforming growth factor beta 1 gene (*TGFB1*, rs1800472 C>T) in the donor (**A – C**, blue line) and the recipient (**D – F**, yellow line).

Next, the donor-recipient pairs were divided into four groups based on the presence or absence of the T-allele in the donor and recipient. Kaplan–Meier survival analyses revealed a significant difference in graft survival among the four groups (Figure 3; *P* = 0.034). Moreover, the T-allele of the *TGFB1* polymorphism in the donor seemed to have a bigger impact on graft survival than the T-allele in the recipient. Recipients with a CT-genotype receiving a graft with the CT-genotype appeared to have the worst outcome. However, this combined genotype was only identified in five donor-recipient pairs.

**Figure 3.**
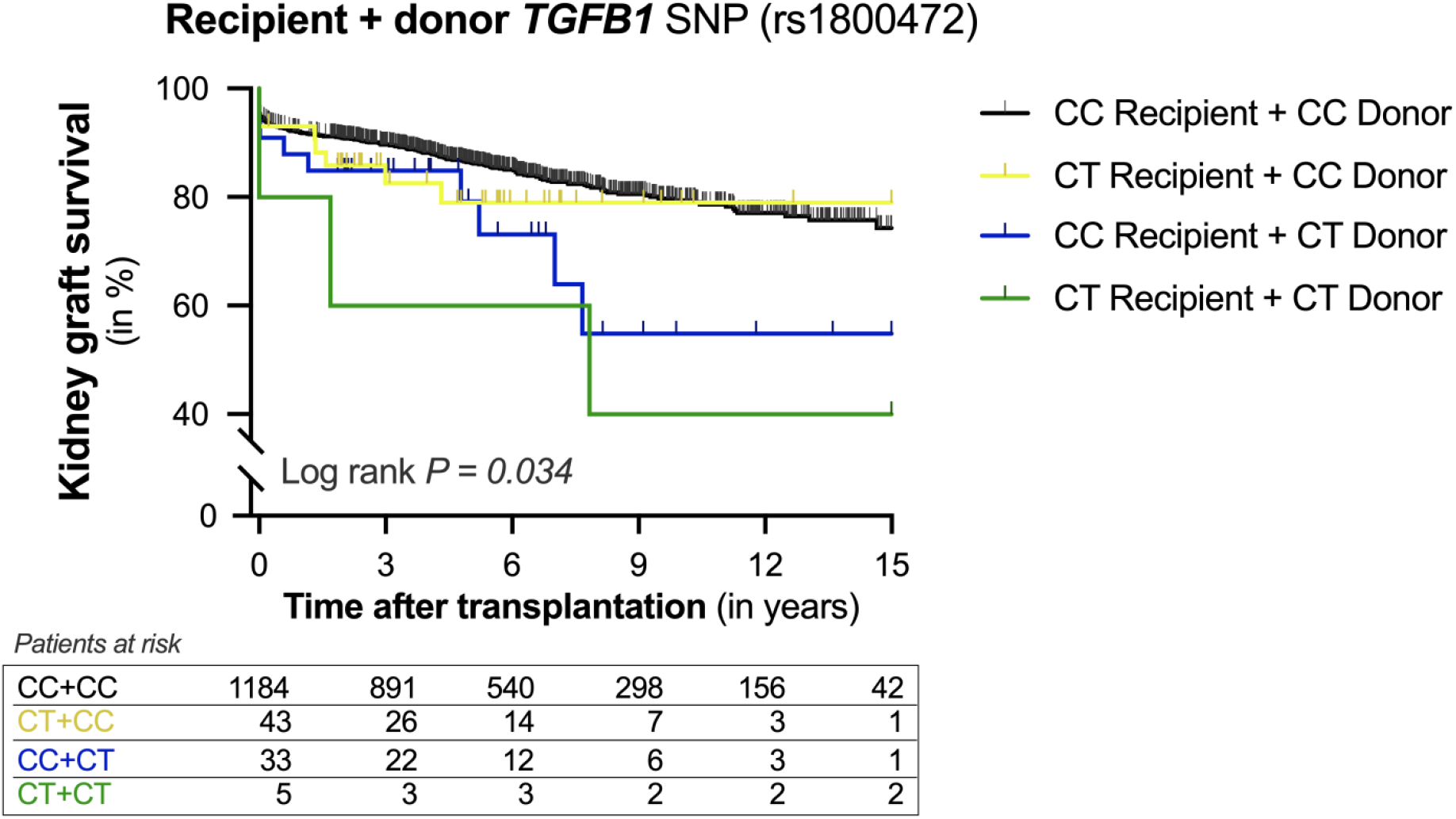
Kaplan-Meier curves for 15-year death-censored graft survival after kidney transplantation according to the presence of the *TGFB1* variant in the donor and recipient. Cumulative 15-year death-censored kidney graft survival according to the presence of the Thr263Ile variant in the transforming growth factor beta 1 gene (*TGFB1*, rs1800472 C>T) in donor-recipient pairs. Pairs were divided into four groups according to the absence (black line) or presence of the T-allele in the recipient (yellow line), donor (blue line) or both (green line). Log-rank test was used to compare the incidence of graft loss between the groups.

### Regression analysis for the *TGFB1* polymorphism and graft loss

Finally, we explored whether the *TGFB1* variant in the donor was an independent risk factor for graft loss. In univariate analysis, the T-allele of the *TGFB1* SNP in the donor was associated with a hazard ratio of 2.12 (95%-CI: 1.18 – 3.79; *P* = 0.012) for graft loss after complete follow-up. Next, multivariable analysis was performed to adjust for potential confounders, including donor and recipient characteristics, and transplant variables (Table 2). In Cox regression analysis, the *TGFB1* SNP in the donor remained significantly associated with graft loss independent of potential confounders. Finally, we performed a multivariable analysis with a stepwise forward selection procedure using all variables that were significantly associated with graft loss in univariable analysis (Table 3). In the final model, the *TGFB1* SNP in the donor, donor and recipient age, recipient blood type, and DGF were included. After adjustment, the T-allele *TGFB1* SNP in the donor was significantly associated with graft loss with a hazard ratio of 2.04 (95% CI: 1.14 – 3.68, *P* = 0.017). Altogether, these results demonstrate that the minor allele of the *TGFB1* variant in the donor associates with a higher risk of graft loss after kidney transplantation.

**Table 2.** Associations of *TGFB1* polymorphism in the donor with graft loss after kidney transplantation.

|  | *TGFB1* SNP (rs1800472-T) in the donor |  |  |
| --- | --- | --- | --- |
|  | Hazard ratio (CT vs CC) | 95% CI | P-value |
| **Model 1** | 2.12 | 1.18 – 3.79 | 0.012 |
| **Model 2** | 2.05 | 1.11 – 3.79 | 0.023 |
| **Model 3** | 2.34 | 1.31 – 4.21 | 0.004 |
| **Model 4** | 2.11 | 1.17 – 3.79 | 0.013 |

Data are presented as a hazard ratio with 95% confidence interval (CI) and *P*-value.
**Model 1:** Crude model.
**Model 2:** Adjusted for model 1 plus recipient characteristics: recipient age, recipient sex, recipient blood type and dialysis vintage.
**Model 3:** Adjusted for model 1 plus donor characteristics: donor age, donor sex, donor blood type, and donor origin.
**Model 4:** Adjusted for model 1 plus transplant characteristics: cold and warm ischemia time, and the occurrence of delayed graft function (DGF).

**Table 3.** **Competitive analysis of the associations of characteristics with graft loss after kidney transplantation**.

| Variables not in the equation |  | Variables in the equation |  |  |
| --- | --- | --- | --- | --- |
| Variables | P-value | Variables | P-value | Hazard Ratio |
| Cold ischemia time<br>(in hours) | 0.054 | rs1800472-T in the donor<br>(CT versus CC) | 0.017 | 2.04 (1.14 – 3.68) |
| Warm ischemia time<br>(in minutes) | 0.07 | Donor age<br>(in years) | 0.003 | 1.02 (1.01 – 1.03) |
| Donor type<br>(living versus deceased) | 0.08 | Recipient blood type<br>(ABO versus other) | 0.001 |  |
| Corticosteroids | 0.12 | Recipient age<br>(in years) | <0.001 | 0.98 (0.97 – 0.99) |
| Cyclosporin | 0.32 | Delayed graft function<br>(yes versus no) | <0.001 | 4.01 (3.03 – 5.31) |
| Donor blood type<br>(ABO versus other) | 0.98 |  |  |  |

Multivariable cox regression was performed for kidney graft survival with a stepwise forward selection. Only variables that with a *P*-value below 0.05 in the univariate analysis were included. Data are presented as a hazard ratio with a 95% confidence interval (CI) and *P*-value. In the final model, the *TGFB1* SNP (rs1800472-T) in the donor, donor age, recipient blood type, recipient age, and the occurrence of delayed graft function were included, whereas cold ischemia time, warm ischemia time, donor type, use of corticosteroids, use of cyclosporin, and donor blood type were not.

## Discussion

New therapeutic strategies to improve long-term allograft survival are urgently needed, but the development of new drugs for kidney transplantation is limited.(26) Studies of human genetics are therefore needed to predict the success of novel drug targets since genetically supported drug targets are more than twice as likely to be successful in clinical trials and lead to approved therapeutics.(27,28) Here, we studied a common functional polymorphism in *TGFB1* to dissect the role of TGF-β1 signaling in kidney transplant survival. The key finding in our study is that kidney allografts possessing a low-producing *TGBF1* polymorphism are associated with a higher risk of graft loss. In contrast, no association was seen between this *TGBF1* polymorphism in the recipient and long-term allograft survival. In conclusion, our study provides genetic evidence that the TGF-β1 pathway in the donor could be favorable for long-term graft survival in kidney transplantation.

Whole-genome sequencing of plasma TGF-β1 recently highlighted the *TGFB1* polymorphism rs1800472 as the top functional variant in a genome-wide association study (GWAS) for plasma TGF-β1 levels using whole-genome sequencing.(21) Furthermore, the overall heritability of the TGF-β1 concentration in plasma was ∼23%, of which 71% of the genetic variance was explained by this polymorphism alone. To our knowledge, our study is the first to demonstrate an association between the *TGFB1* Thr263Ile variant in the donor and long-term graft survival after kidney transplantation. In particular, we found that the T-allele in the donor approximately doubled the risk of graft loss. Previously, the minor alleles of two other *TGFB1* polymorphisms (rs1800470-C>T and rs1800471-G>C) in the donor have also been associated with worse graft survival after kidney transplantation.(29) In line with our results, the minor alleles of these *TGFB1* polymorphisms have also been suggested to lead to lower levels of TGF-β1.(30–32) Furthermore, Du et al. reported that long-term survival kidney transplant recipients had higher TGF-β1 levels than short-term survival kidney transplant recipients.(33) Serum TGF-β1 levels positively correlated with long-term graft survival and function. Altogether, our study adds to a growing body of evidence that indicates that TGF-β1 has protective effects on kidney transplant survival.

The impact of the recipient *TGFB1* genotype on outcome after kidney transplantation has been investigated by various studies but remains controversial.(34–38) A recent metanalysis of nine studies including 352 rejection cases and 882 controls concluded that the recipient TGFB1 genotype was not significantly associated with acute rejection after kidney transplantation.(35) Similarly, we also found no association between the *TGFB1* polymorphism rs1800472 in the recipient and BPAR (Supplementary Data). In the past, low-producing genotypes of *TGFB1* in the recipient have been associated with both superior and worse outcomes in kidney transplantation,(34,37) while others have found no association with kidney allograft survival.(36) In our transplant cohort of 1,271 donor-recipient pairs, we did not find an association between a low-producing *TGBF1* polymorphism in the recipient and graft survival either. We also assessed the relationship between the *TGFB1* polymorphism rs1800472 and DGF. While we did see a trend for higher risk of DGF in recipients carrying the T-allele of the *TGFB1* polymorphism, we did not find an association between the donor *TGFB1* genotype and DGF. Similar to our observations the study by Israni and colleagues found no association for the *TGFB1* polymorphism rs1800472 in the donor and DGF.(5) In conclusion, our results suggest that it is not the circulating TGF-β1 from the recipient, but rather the local TGF-β1 expression by the donor kidney that promotes graft survival in kidney transplantation.

Generally, TGF-β1 is considered to be a critical driver of fibrosis.(10) Given the abundance of evidence from animal models and translational studies, inhibiting the TGF-β1 signaling pathways would hypothetically prevent the development of kidney fibrosis in kidney disease and transplantation.(39,40) However, contrary to expectations, results from clinical trials have been underwhelming, as therapies targeting TGF-β1 have not translated into approved treatment for patients.(8,41,42) Emerging data demonstrate that TGF-β1 is not only capable of inducing fibrosis, but also has protective effects.(7) In conformity with the findings, loss-of-function mutations in the *TGFB1* gene were recently shown to cause severe inflammatory bowel disease and encephalopathy in humans, demonstrating the anti-inflammatory properties of this cytokine.(43) In preclinical transplantation studies, TGF-β1 was shown to protect against brain death-induced organ damage, ischemia-reperfusion injury, and prolong graft survival.(44–47) The mechanisms behind these protective effects include (i) Protecting kidney cells against apoptosis, (ii) Stimulating tissue regeneration, and (iii) Diminishing alloimmunity to kidney transplants by inducing tolerance through regulatory T cells.(12)

Several limitations of our study warrant consideration. First and foremost, our study is observational in nature and can therefore not prove causality. Further studies are needed to assess whether the observed association is indeed causal. Furthermore, we examined one polymorphism in *TGFB1* and did not assess *TGFB1* haplotypes. Lastly, the relationship between genotypes and plasma levels of TGF-β1 was not assessed in our cohort due to the lack of samples. In contrast, crucial strengths of the current study include the analysis of a functional polymorphism in both the donor and recipient, the large sample size, and the stringent and clinically meaningful endpoint, and the lengthy follow-up time.

In conclusion, we found that patients receiving a donor kidney carrying the T-allele of the *TGFB1* polymorphism rs1800472 have a higher risk of late graft loss. Considering that this T-allele is a low-producing *TGBF1* variant, our findings imply a beneficial effect of TGF-β1 signaling on long-term allograft survival in kidney transplantation.

## Data Availability

Data is available upon request.

## Abbreviations

BPAR: Biopsy-proven acute rejection
CIT: Cold ischemia time
DBD: Donation after circulatory death
DCD: Donation after brain death
DGF: Delayed graft function
ESKD: End-stage kidney disease
HLA: Human leukocyte antigen
HR: Hazard ratio
PRA: Panel-reactive antibody
SMAD: Small mothers against decapentaplegic homologue
SNP: Single-nucleotide polymorphism
TGF-β: Transforming growth factor beta
TGF-β1: Transforming growth factor beta 1
*TGFB1*: Transforming growth factor beta 1 gene
WIT: Warm ischemia time

## Acknowledgment

The authors thank the members of the REGaTTA cohort (REnal GeneTics TrAnsplantation; University Medical Center Groningen, University of Groningen, Groningen, the Netherlands): S. J. L. Bakker, J. van den Born, M. H. de Borst, H. van Goor, J. L. Hillebrands, B. G. Hepkema, G. J. Navis and H. Snieder. The illustrations of Figure 1 were made by Siawosh K. Eskandari.

## Supplementary Data

**Table S1.**
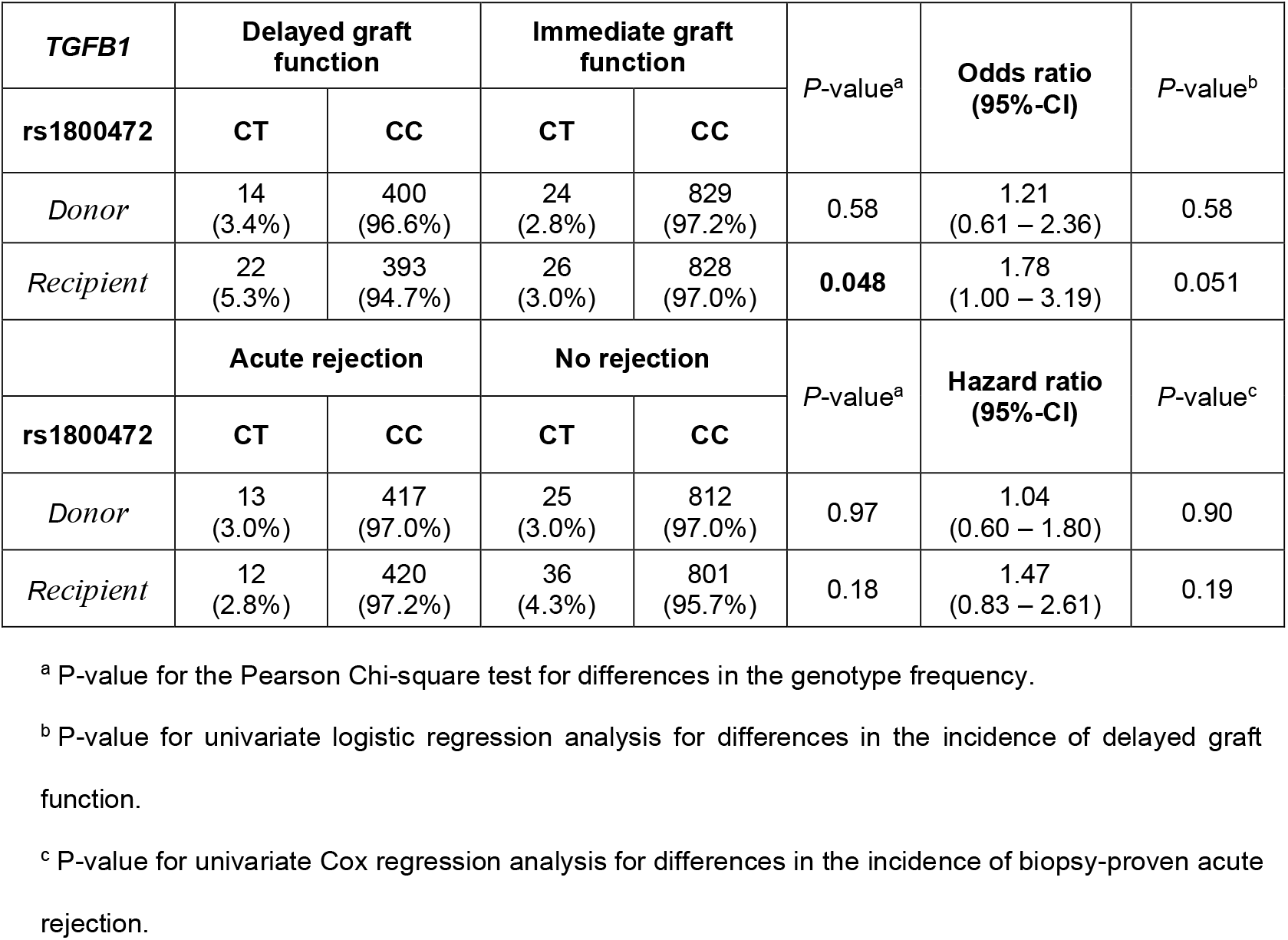
Genotype frequency and hazard ratios for delayed graft function and acute biopsy-proven rejection for TGBF1 genotypes.

| *TGBF1* | Delayed graft function |  | Immediate graft function |  | *P*-value<sup>a</sup> | Odds ratio (95%-CI) | *P*-value<sup>b</sup> |
| --- | --- | --- | --- | --- | --- | --- | --- |
| **rs1800472** | **CT** | **CC** | **CT** | **CC** |  |  |  |
| *Donor* | 14<br>(3.4%) | 400<br>(96.6%) | 24<br>(2.8%) | 829<br>(97.2%) | 0.58 | 1.21<br>(0.61 – 2.36) | 0.58 |
| *Recipient* | 22<br>(5.3%) | 393<br>(94.7%) | 26<br>(3.0%) | 828<br>(97.0%) | **0.048** | 1.78<br>(1.00 – 3.19) | 0.051 |
|  | Acute rejection |  | No rejection |  | *P*-value<sup>a</sup> | Hazard ratio (95%-CI) | *P*-value<sup>c</sup> |
| **rs1800472** | **CT** | **CC** | **CT** | **CC** |  |  |  |
| *Donor* | 13<br>(3.0%) | 417<br>(97.0%) | 25<br>(3.0%) | 812<br>(97.0%) | 0.97 | 1.04<br>(0.60 – 1.80) | 0.90 |
| *Recipient* | 12<br>(2.8%) | 420<br>(97.2%) | 36<br>(4.3%) | 801<br>(95.7%) | 0.18 | 1.47<br>(0.83 – 2.61) | 0.19 |

<sup>a</sup> *P*-value for the Pearson Chi-square test for differences in the genotype frequency.
<sup>b</sup> *P*-value for univariate logistic regression analysis for differences in the incidence of delayed graft function.
<sup>c</sup> *P*-value for univariate Cox regression analysis for differences in the incidence of biopsy-proven acute rejection.

**Table S2.** Subgroup analysis for the association between graft loss and recipient TGBF1 genotypes.

| Recipient<br>rs1800472<br>*TGFB1* | Functioning graft |  | Graft loss |  | *P*-value<sup>a</sup> | Hazard ratio<br>(95%-CI) | *P*-value<sup>b</sup> |
| --- | --- | --- | --- | --- | --- | --- | --- |
|  | CT | CC | CT | CC |  |  |  |
| *Female recipient* | 12<br>(2.7%) | 435<br>(97.3%) | 5<br>(6.0%) | 78<br>(94.0%) | 0.11 | 2.19<br>(0.80 – 6.03) | 0.13 |
| *Male recipient* | 25<br>(4.1%) | 582<br>(95.9%) | 6<br>(4.5%) | 126<br>(95.5%) | 0.83 | 1.60<br>(0.70 – 3.65) | 0.27 |
| Recipient<br>rs1800472<br>*TGFB1* | Functioning graft |  | Graft loss |  | *P*-value<sup>a</sup> | Hazard ratio<br>(95%-CI) | *P*-value<sup>b</sup> |
|  | CT | CC | CT | CC |  |  |  |
| *Deceased kidney donor* | 31<br>(3.9%) | 766<br>(96.1%) | 9<br>(4.7%) | 181<br>(95.3%) | 0.60 | 1.67<br>(0.85 – 3.28) | 0.14 |
| *Living kidney donor* | 6<br>(2.3%) | 251<br>(97.7%) | 2<br>(8.0%) | 23<br>(92.0%) | 0.10 | 2.32<br>(0.31 – 17.49) | 0.41 |

Subgroup analyses was performed for recipient sex as well as donor type for graft loss according to the presence of the *TGFB1* rs1800472 polymorphism in the recipient.
<sup>a</sup> *P*-value for the Pearson Chi-square test for differences in the genotype frequency.
<sup>b</sup> *P*-value for univariate Cox regression analysis for differences in the incidence of graft loss after complete follow-up.

